# The Impact of Environmental Risk Factors on Delirium and Benefits of Noise and Light Modifications: A Scoping Review

**DOI:** 10.1101/2020.05.20.20108373

**Authors:** Haleh Hashemighouchani, Julie Cupka, Jessica Lipori, Matthew M Ruppert, Elizabeth Ingersent, Tezcan Ozrazgat-Baslanti, Parisa Rashidi, Azra Bihorac

## Abstract

**Purpose:** To explore existing literature on the association between environmental risk factors with delirium and to investigate the effectiveness of environmental modifications on prevention or management of delirium.

**Materials and Methods:** This is a scoping review of peer-reviewed studies in Pubmed and the reference lists of reviewed articles. Observational studies reporting the effect of noise, light, and circadian rhythm on delirium and interventional studies assessing delirium in modified environments were reviewed.

**Results:** Thirty eight studies were included, of which, 21 evaluated impact of environment on delirium, and 16 studied the interventions. Interventions targeted reducing noise exposure, improving light exposure to follow circadian rhythm, and promoting sleep. Mixed findings of the reviewed studies yielded to inconclusive results; however quiet-time protocols, earplugs, and bright light therapy might benefit prevention, or management of delirium.

**Results:** Thirty seven studies were included, 21 of which evaluated the impact of environment on delirium and 16 studied possible solutions to mitigate those impacts. Mixed findings of the reviewed studies yielded inconclusive results; a clearly delineated association between high noise levels, abnormal amounts of light exposure, and sleep disruption with delirium could not be established. Interventions targeted reducing noise exposure, improving day-time and mitigating night-time light exposure to follow circadian rhythm, and promoting sleep. The overall evidence supporting effectiveness of environmental interventions was also of a low confidence; however, quiet-time protocols, earplugs, and bright light therapy showed a benefit for prevention or management of delirium.

**Conclusion:** Environmental modifications are non-invasive, risk-free, and low-cost strategies that may be beneficial in preventing and managing delirium, especially when used as part of a multi-component plan. However, given the limited evidence-based conclusions, further high-quality and larger studies focusing on environmental modifications and delirium outcomes are strongly recommended.

## INTRODUCTION

Delirium is a multifactorial, acute, state of confusion characterized by disturbance of consciousness and cognition; it is particularly common in the intensive care unit (ICU) with incidence ranging from 19 to 87% with especially higher rates in mechanically ventilated patients ^1–3^. ICU delirium is associated with adverse outcomes, including prolonged mechanical ventilation, increased risk of long term cognitive dysfunction, prolonged ICU and hospital stays, higher cost of care, and increased mortality ^4–7^. While the pathophysiology of delirium is poorly understood, there are multiple factors associated with an increased risk for developing delirium, including age, level of education, pre-existing conditions such as hypertension, neurological or psychological disorders, illness severity, Acute Physiology and Chronic Health Evaluation II (APACHE II) score, sensory impairment, and use of analgesics, sedatives, and polypharmacy ^8–12^. In addition, the ICU environment may contribute as a potentially modifiable risk factor for developing delirium. Decreased natural daylight, night-time light exposure, excessive noise, immobilization, use of physical restraints, and isolation are potential delirium risk factors in ICU ^13–15^.

ICU noise levels are above internationally recommended levels by the World Health Organization’s (WHO) recommendations, which suggest 30 A-weighted decibels (dBA) for background noise, a maximum of 35 dBA for treatment and observation areas, and a maximum of 40 dBA at night in a hospital setting ^16–18^. Patients interviewed post-ICU discharge report noise as an overall stressor and contributor to loss of sleep ^19, 20^. Another common environmental disturbance for ICUs is non-cycling light sources. Disruptions in normal amounts of blue light (460-480 nm) hitting the retina affect neurological processes responsible for melatonin release ^15^. Constant delivery of these wavelengths may cause abnormal suppression of melatonin release, thus potentially, altering circadian cycles ^15^. Although the nature of the ICU environment does not lend itself to quietude, it is feasible to employ noise-reducing and light modifications that synchronize circadian rhythm, facilitating recovery.

Prevalence of delirium-associated adverse effects and the multitude of risk factors in the ICU make delirium prevention and management essential. Current strategies include pharmacological, non-pharmacological, and multi-component interventions geared towards decreasing delirium incidence and duration. Pharmacological interventions focus on haloperidol and dexmedetomidine, with limited research into ramelteon, melatonin, and ziprasidone ^21–24^ The largest clinical trial to date on haloperidol or ziprasidone in delirious patients failed to show significant clinical benefit ^23^, and current literature does not support use of anti-psychotic agents, benzodiazepines, or melatonin in delirium management ^21, 25^. Given the lack of evidence supporting pharmacological measures, research into efficacy of non-pharmacological techniques such as noise reduction or dynamic lighting is crucial. Implementing effective delirium management strategies shows promise in decreasing morbidity, mortality, length of stay, and resource burden in the ICU ^2^. The purpose of this scoping review is to map existing literature identifying modifiable environmental risk factors for delirium, and assess the role of non-pharmacological noise, light, and circadian rhythm interventions for delirium prevention and management.

## MATERIALS AND METHODS

This review was conducted according to methods of Arksey and O’Malley ^26^ and Levac et al. ^27^, and reported following the Preferred Reporting Items for Systematic Reviews and Meta-Analyses (PRISMA) Extension for Scoping Reviews ^28^ (Supplementary Table 1). The aim of this review is to examine the extent and nature of available literature, and highlight areas requiring further inquiry regarding these questions: “How do environmental noise, light, and disrupted circadian rhythms affect delirium?” and “How do existing environmental interventions help prevent or manage delirium?”

### Search Strategy and data charting

Studies were identified by searching Pubmed for articles relating to our questions. Search results were restricted to the English language and peer-reviewed studies, with no restriction on year of publication. Search queries were generated using the following combination of keywords: [“delirium” AND “noise OR sound OR light OR circadian”]. The search was applied with no field tags to maximize results.

After compiling research results and removing duplicates, two authors screened titles and abstracts to retrieve articles for eligibility. Articles on pediatric populations, animal subjects, case reports, or if the full-text was unavailable were excluded. Additional studies were identified through hand-searches and searching the reference list of reviewed articles. Three authors reviewed the full text of eligible articles and extracted data using a pre-designed worksheet reviewed and tested by the team before data charting (Supplementary Table 2). Elements of the data charting worksheet included study design, setting, sample size, aim, detailed methodology, characteristics of intervention and control groups, measured outcomes, diagnostic tools, main conclusions, and study strengths and limitations. Any disagreements were resolved by thoroughly discussing any points of concern.

We included observational studies analyzing association between noise levels, light exposure, or disrupted circadian rhythm with delirium, and interventional studies assessing effectiveness of modified noise or light exposure or improved circadian rhythm on delirium. Articles were excluded if environmental intervention was an element of a multi-component non-pharmacological bundle, not the main focus. In initial full text review and data charting, we reviewed all interventional articles reporting results on delirium or environmental risk factors of delirium, including noise or light levels, and quality/quantity of sleep. We acknowledge these outcomes are modifiable risk factors linked to delirium prevention or management; however, to better map the existing literature related to our research question, we excluded articles without results linked to delirium. These articles are in Supplementary Table 3.

## RESULTS

### Literature search results & outcome

The electronic database search retrieved 457 articles, which were screened by title and abstract, resulting in 166 studies for full-text review. Hand-search and searching reference lists added 28 additional articles. During full-text review of these 194 articles, 157 were excluded. In total 37 studies were included: 21 assessed association between environmental risk factors and delirium ^13, 19, 20, 29–46^, and 16 reported on delirium after an environmental intervention ^7 14, 15, 18, 47–58^ (Figure 1).

**Figure 1.**
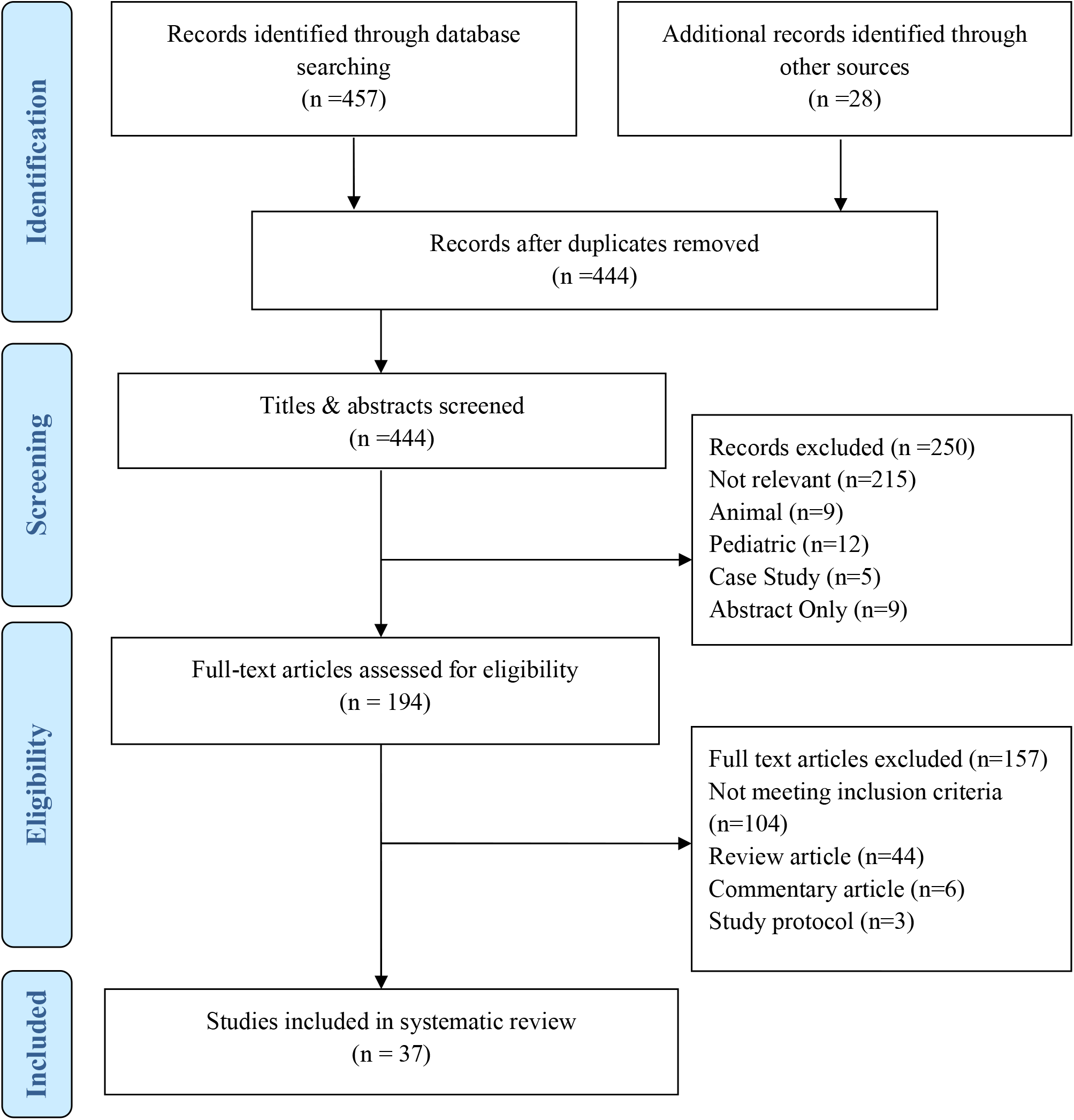
PRISMA Record Screening Flow chart.

### Characteristics of the reviewed articles

Included studies were conducted between 1997 to 2019, in the USA ^19, 30, 36, 38, 42, 45, 57, 58^, the Netherlands ^14, 31, 47, 49, 50^, Japan ^37, 40, 51, 52^, France ^32, 35, 56^, Belgium ^13, 48^, Denmark ^15, 33^, Italy ^41, 46^, Sweden ^18, 20^, Canada ^34^, China ^44^, India ^39^, Israel ^29^, Singapore ^54^, South Korea ^53^, Thailand ^55^, Turkey ^43^, and UK ^7^. Thirty-one studies were conducted among critically ill patients while five reviewed general hospital populations ^29, 34, 40, 43, 53^, and one a geriatric monitoring unit for acute delirium care ^54^. Among the 37 reviewed articles, all observational association studies and 12 interventional studies reported delirium incidence, while 2 interventional studies measured delirium prevalence ^18, 57^. Delirium severity was assessed in three of the interventional studies ^47, 53, 54^. Three articles also reviewed delirium duration ^7, 47, 50^.

Most studies assessed delirium using Confusion Assessment Method for the ICU (CAM-ICU) ^7, 15, 18, 19, 33, 35, 36, 38, 39, 41, 44–47, 50, 55–58^; other identification methods included validated Dutch CAM-ICU ^31^, Confusion Assessment Method (CAM) ^40, 49, 54^, Intensive Care Delirium Screening Checklist (ICDSC) ^14, 32, 42^, Neelon and Champagne Confusion Scale (NEECHAM) ^13 48^, non-validated ^51^ and validated ^52^ Japanese NEECHAM, Delirium Observation Screening Scale (DOSS) ^49^, behavioral observations based on the Diagnostic and Statistical Manual of Mental Disorders, 3^rd^ edition (DSM-III) ^34^, 3^rd^ edition-revised (DSM-III-R) ^37^, and 4^th^ Edition (DSM-IV) ^20, 40, 43^, and behavioral observations based on International Classification of Diseases, 9^th^ Revision, Clinical Modification (ICD-9-CM) criteria ^29^. One study used both retrospective chart review and site-specific pre-specified criteria based on new and rapid onset of disturbed consciousness and/or perceptual disturbances ^30^. Studies assessed delirium severity by non validated Delirium Severity Index (DSI) ^47^, Delirium Rating Scale (DRS) ^53^, Delirium Rating Scale-Revised-98 (DRS-R-98) ^54^, and Memorial Delirium Assessment Scale (MDAS) ^53^. Specific study details including study design, setting, sample size, method details, outcomes, and a brief description of findings with statistics are summarized in Table 1 for observational studies reporting on environmental risk factors, and Table 2 for environmental intervention studies.

**Table 1:** Summary of Characteristics and Findings of Reviewed Observational Association Studies.

| Study<br>(Author, Year,<br>Country) | Study Design | Setting & Population<br>Characteristics | Examined Risk<br>Factors | Method Details | Findings | Statistics |
| --- | --- | --- | --- | --- | --- | --- |
| **Noise** |  |  |  |  |  |  |
| Johansson,<br>2012, Sweden <sup>20</sup> | Observational,<br>pre-study | ICU (general medical-<br>surgical)<br>n=13<br>ICU patients excluding head<br>injury, hearing impairment,<br>dementia | A-weighted<br>decibel<br>measurements<br>Post-ICU<br>survey on<br>memories of<br>ICU<br>environment | Delirium: hourly behavioral<br>observations by nurse based on<br>Granberg-Axell protocol (2001) and<br>DSM-IV<br>Noise: Bruel & Kjaer 2260 sound<br>level meter placed close to patient<br>bed, one-minute average interval of<br>A-weighted sound levels analyzed<br>with B&K Evaluator software<br>Memory survey: open-ended,<br>unstructured interview after ICU<br>discharge focusing on memories of<br>ICU environment and sounds | No association between<br>high number of early<br>signs of ICU delirium<br>and high sound levels<br>Interview responses:<br>mixed, some sound<br>memories were<br>scary/irritating, others<br>were comforting,<br>unnoticed, or<br>incorporated into<br>dreams | p > 0.05 for all sound<br>and delirium analyses<br>No statistics reported<br>for interviews. |
| Knauert, 2016,<br>USA <sup>19</sup> | Observational,<br>prospective | MICU<br>n=59<br>All adult patients admitted<br>within 48 hours before next<br>sound recording period,<br>excluding those expected to<br>die within 24 hours,<br>undergoing comfort care, or<br>expected to be transferred<br>before study completion | Leq &<br>frequency of<br>peak<br>occurrences | Delirium: CAM-ICU daily<br>Noise: two Extech HD600 sound<br>meters placed at foot of patient bed<br>with standardized distance from<br>care equipment, 10-second interval<br>of A- and C-weighted sound levels,<br>decibel range set to 30-130 dB,<br>detector response set to 'fast, 125<br>milliseconds' | Delirium was not<br>associated with Leq or<br>peak occurrences | p > 0.05 for all sound<br>and delirium analyses |
| **Light** |  |  |  |  |  |  |
| Balan, 2001,<br>Israel <sup>29</sup> | Observational,<br>retrospective | Geriatric hospital<br>n=234<br>Patients aged ≥65 years and<br>older, excluding patients<br>admitted with pre-existing<br>delirium or those unable to<br>communicate due to<br>cognitive impairment | Season of<br>delirium<br>diagnosis | Delirium: ICD-9-CM criteria,<br>assessed and diagnosed by a<br>psychiatrist after development of<br>any abrupt change in mental or<br>behavioral condition<br>Light: patients compared by season<br>of admission (winter, December-February; spring, March-May;<br>summer, June-August; fall,<br>September-November) | Significantly higher<br>rates of delirium in<br>winter than summer<br>months | X<sup>2</sup>, 2 df = 14.36<br>p < 0.001 |
| Kohn, 2013, USA <sup>30</sup> | Observational, retrospective | MICU<br>n= 6336<br>All admitted patients, restricted to the index ICU admission during a hospitalization<br>SICU<br>n = 6660<br>All admitted patients, restricted to the index ICU admission during a hospitalization | Presence of a window<br>Whether view out window was a natural or industrial view<br>Presence of a half- or full-sized window for windows facing the same direction | Delirium, MICU: retrospective chart review of random patient sample (7%); diagnosed if specific keywords associated with delirium were documented by physician or nurse on at least 2 separate days<br>Delirium, SICU: screened daily by nurse practitioner for pre-specified criteria based on new and rapid onset of disturbed consciousness and/or perceptual disturbance<br>Windows, both units: whether patient was admitted to room with or without a window; whether window had a natural or industrial view; whether window was half- or full-sized | No association with delirium incidence and the presence of a window, in all analyses<br>No association with delirium incidence and a natural or industrial view, in all analyses<br>No association between delirium incidence and presence of a half- or full-sized window for windows facing the same direction, in all analyses | p > 0.05 for all light and delirium analyses |
| Simons, 2014, Netherlands <sup>31</sup> | Observational, retrospective | ICU<br>n=3198<br>All patients who were admitted to the ICU within 30 days of hospital admission, restricted to the first ICU admission during a hospitalization | 7-, 28-, and 60-day prehospital photoperiod<br>Season of admission<br>Subgroup analysis: 28-day photoperiod in patients admitted to ICU within 48 hours of hospital admission | Delirium: Dutch validated CAM-ICU at least twice daily during complete ICU stay<br>Light: sunlight data was obtained from nearby weather stations of the Royal Dutch Meteorological Institute; cumulative photoperiod was calculated from the total amount of radiation, defined as total number of hours of daylight for 7, 28, and 60 days before hospital admission | No association between delirium incidence and prehospital sunlight exposure for all photoperiods (7-, 28-, 60-day)<br>No association between delirium incidence and season of admission<br>Subgroup analysis: no association between 28-day photoperiod and delirium incidence | p > 0.05 for all light, season, and delirium analyses |
| Smonig, 2019, France <sup>32</sup> | Observational, prospective | MICU<br>n=195<br>Consecutive adult patients requiring invasive MV in the ICU for at least 2 days, without acute brain injury or<br>conditions interfering with delirium assessment (i.e. dementia, deaf, blind) | Presence of a window | Delirium: RASS followed by ICDSC twice daily, defined as the presence of ICDSC $\geq 4$ for at least 2 consecutive ICU days<br>Light: exposure determined by whether patient was assigned to a room with or without windows | No association between exposure to windows and delirium burden (incidence and duration), even when excluding room transfers | p > 0.05 for all light and delirium analyses |
| van Rompaey, 2009, Belgium<sup>13</sup> | Observational, prospective | ICU (mixed)<br>n=523<br>All consecutive patients aged $\geq 18$ years with ICU stay of $\geq 24$ hours; patients were enrolled when GCS reached at least 10. | Absence of visible daylight | Delirium: NEECHAM Confusion Scale (frequency not specified)<br>Light: exposure determined by whether patient was exposed to visible daylight during ICU stay | Patients had a significantly higher risk of developing delirium with the absence of visible daylight | Univariate<br>OR 1.75<br>95% CI (1.19-2.56)<br>p = 0.003<br>Multivariate<br>OR 2.39<br>95% CI (1.28-4.45) |
| **Sleep** |  |  |  |  |  |  |
| Boesen, 2016, Denmark<sup>33</sup> | Observational, prospective descriptive | ICU (mixed)<br>n=14<br>Mechanically ventilated patients $\geq 18$ years, without structural neurological illnesses or administration of propofol or benzodiazepines | PSG results<br>Sleep as recorded by CBO | Delirium: SAS & CAM-ICU once per shift<br>Sleep, PSG: 24 hour simplified PSG with a 2-lead-frontal EEG (no occipital or central leads), 2-lead EOG, 1 chin EMG, and 1-lead ECG; G&B Electronic Designs Ltd: Lifelines, TrackIT screener PP3-S; recording started at noon; PSGs were scored by an EEG technician in 30 second epochs according to the AASM standards; atypical PSGs were re-scored; due to encephalopathy, wakefulness was interpreted using eye-blinking and EEG reactivity<br>Sleep, CBO: 24 hour clinical sleep registration done by attending nurses, noted on a case report form as “asleep” or “awake”; awakenings were registered as transitions from “asleep” to “awake”; measurements included total clinical time awake, total clinical time asleep, and number of hours with logged entries | No clear differences in sleep patterns between patients with and without delirium, for both PSG and CBO analysis<br>The more pathological the EEGs and the patients were, the less correlation with clinically observed sleep was seen<br>Sleep quality and quantity cannot be feasibly assessed with PSG in mechanically ventilated ICU patients, since the vast majority of PSGs were atypical without objective sleep signs | p > 0.05 for all sleep and delirium analyses |
| Bowman, 1997, Canada<sup>34</sup> | Observational, descriptive | Teaching hospital<br>n=43<br>Elderly subjects undergoing orthopedic hip surgery (age | Sleep satisfaction | Delirium: DSM-III diagnosis as evidenced by notes in RN reports, chart review, interview with on-duty RNs, or assessment by researcher during researcher’s daily rounds; | Patients who developed post-operative delirium had poorer sleep satisfaction than those without post-operative | Day 2<br>p = 0.008574<br>Day 3<br>p = 0.031772 |
| | | criteria not specified); subjects with known dementia or MMSE score $\leq 23$ were excluded | | MMSE repeated daily until subject obtained score of $\geq 24$<br>Sleep: previous night's sleep satisfaction recorded every AM for 5 days using a seven-point Likert scale | delirium, except postoperative day 5 | $p > 0.05$ for all other sleep and delirium analyses |
| Drouot, 2012, France <sup>35</sup> | Observational, retrospective | MICU<br>n=57<br>Adult, conscious, non-sedated ICU patients admitted for acute respiratory failure & treated with NIV for at least 2 days, without presence of: CNS disorder, GCS $< 15$, delirium and/or confusion, administration in past 2 days of drugs interfering with sleep & EEG activity | PSG results | Delirium: CAM-ICU daily<br>Sleep: Embla S700 digital recorder from 1500 to 0800 on the following day; leads included: three EEG channels, chin EMG, two EOGs, submental EMG, two tibialis anterior EMGs, and ECG; EEG signals amplified and recorded at 200-Hz sampling frequency, filtered (0.5–70 Hz); Rechtschaffen and Kales criteria were used to score sleep stages and awakenings | Significant association between delirium occurrence and atypical sleep on EEG | $p < 0.05$ |
| Kamdar, 2015, USA <sup>36</sup> | Secondary analysis of prospective observational study | MICU<br>n=223<br>Patients with $\geq 1$ night in the MICU in between two consecutive days of delirium assessment, data analysis was only on patients' first MICU admission | Perceived sleep quality | Delirium: CAM-ICU twice daily at 0800 and 2000<br>Sleep: RCSQ daily with an additional item to evaluate perceived night-time noise | No association between transition to delirium and perceived sleep quality | $p > 0.05$ for all sleep and delirium analyses |
| Kaneko, 1997, Japan <sup>37</sup> | Observational | HCU<br>n=36<br>Patients aged over 70 years undergoing gastrointestinal surgery | Sleep-cycle disturbances | Delirium: DSM-III-R (frequency not specified)<br>Sleep: clinical behavioral observations on sleep & wakefulness recorded in two blocks (daytime, 0600-1800; night-time 1800-0600) | Postoperative abnormal sleep patterns are significantly associated with development of delirium | $p < 0.05$ |
| Knauert, 2014, USA <sup>38</sup> | Observational, cross-section pilot | MICU<br>n=29 | Atypical sleep on PSG | Delirium: CAM-ICU on day of enrollment and during PSG (frequency not specified) | No significant relationship between delirium incidence and atypical sleep | $p > 0.05$ for all sleep and delirium analyses |
|  |  | Adults admitted to the MICU for less than 72 hours without terminal illness, coma or deep sedation, severe agitation, or anatomic contraindications to PSG evaluation |  | Sleep: unattended PSG for up to 24 hours via Compumedics' Safiro Portable Data Acquisition System; initiated in the evening; leads included: 6 EEG channels, chin EMG, right and left EOG; ECG; 200 Hz sampling frequency and filtered (0.5 - 70 Hz); Compumedics' Profusion 2 software; if no stage could be marked, the epoch was marked 'unsure'; if epoch could be identified as sleep, but stage was unclear, it was marked 'sleep (no stage)' |  |  |
| Kumar, 2017, India <sup>39</sup> | Observational, pilot prospective derivation cohort | SICU (cardiac)<br>n=120<br>consecutive adult patients undergoing elective or emergency cardiac surgery without preexisting delirium or deafness | Sleep deprivation | Delirium: assessed once daily with RASS followed by CAM-ICU starting on day of extubation<br>Sleep: method of measurement not specified | Sleep deprivation was not significantly related to delirium | p > 0.05 for all sleep and delirium analyses |
| Shigeta, 2001, Japan <sup>40</sup> | Observational | General hospital<br>n=29<br>Patients undergoing laparotomy for digestive disease | Sleep disturbances | Delirium: CAM followed by DSM-IV diagnosis<br>Sleep: subjects' sleep was monitored every 2 hours for 5 days after surgery | All patients with delirium showed sleep disturbances with reversal of the diurnal sleep cycle, including delayed sleep onset, frequent waking, and increased daytime sleep on the following day | No statistics reported. |
| Trompeo, 2011, Italy <sup>41</sup> | Observational | ICU<br>n=29<br>Patients aged 18-75 with MV for at least 2 days for acute respiratory failure related to surgery, excluding chronic dementia, psychosis, mental retardation, stroke, Alzheimer disease, Parkinson disease, central sleep apnea, drug or alcohol abuse | PSG results | Delirium: CAM-ICU twice daily<br>Sleep: NPB-Mallinckrodt Sandman PSG done from 2200-0800, scored using Rechtschaffen and Kales criteria | Delirium is independently associated with severe REM sleep reduction | p = 0.002 |
| Whitcomb, 2013, USA <sup>42</sup> | Observational, pilot | MICU (pulmonary)<br>n=7<br>65 years or older, intubated & sedated without a diagnosis preventing mental awareness assessment | Sleep stages | Delirium: ICDSC once daily<br>Sleep: Zeo wireless sleep monitor, dry silver-coated fabric headband sensor with single bipolar channel, signal includes EEG/EOG/EMG, captured at 128 samples per second and filtered to a frequency of 2 to 47 Hz, microprocessor reports the sleep stage every 30 seconds in real time via artificial neural network using a reduced set of sleep stages including wakefulness, REM sleep, light sleep (combined stages 1 and 2), and deep sleep (combined stages 3 and 4) | Preliminary results suggest a relationship between lack of REM sleep and delirium | No statistics reported. |
| Yildizeli, 2005, Turkey <sup>43</sup> | Observational, retrospective | General hospital<br>n=432<br>All patients older than 18 years who were admitted for thoracotomy or sternotomy with an expected stay of 2 or more days | Sleep deprivation | Delirium: once delirium symptoms were first noted, a psychiatric consult determined diagnosis based on DSM-IV criteria<br>Sleep: method of measurement not specified | Univariate and multivariate analyses showed a significant association between delirium incidence and sleep deprivation | Univariate<br>p = 0.008<br>Multivariate<br>OR 5.642<br>p = 0.05 |
| Zhang, 2015, China <sup>44</sup> | Observational, prospective cohort | ICU (cardiovascular)<br>n=249<br>adult, post-CABG patients without preoperative diagnoses of delirium, mental disease, or dementia | Quality of sleep | Delirium: assessed three times daily (0800, 1600, 2400) with RASS followed by CAM-ICU, and if patient developed change in mental status<br>Sleep quality: assessed via patient self-report, poor quality was defined by symptoms of sleep disorder or sleep deprivation | Poor sleep quality was the strongest independent predictor of delirium | Univariate<br>p < 0.001<br>Multivariate<br>OR 5.001<br>95% CI (2.476-10.101)<br>p < 0.0001 |
| **Multiple Factors** |  |  |  |  |  |  |
| Davoudi, 2019, USA <sup>45</sup> | Observational, prospective pilot | SICU<br>n=22<br>All adult patients expected to stay in ICU more than 2 day and able to wear an ActiGraph device | Sound pressure levels<br>Light intensity<br>Sleep quality | Delirium: CAM-ICU daily, defined as being delirious throughout study period<br>Noise: iPod with a sound pressure recording application measured in dB, device placed on wall behind patient bed | Average night-time sound pressure levels were significantly higher in delirious patients<br>Light levels were significantly different | Sound<br>p < 0.05<br>Light<br>p < 0.05<br>Ability to fall asleep<br>p = 0.01 |
|  |  |  |  | Light: ActiGraph sensor placed on wall behind patient bed at height of patient head<br>Sleep: Freedman Sleep Questionnaire daily | between delirious and non-delirious patients<br>No statistical association between overall quality of sleep and delirium, but delirious patients were significantly more likely to report difficulty falling asleep and to find lighting disruptive at night | Whether night-time lighting was disruptive<br>p = 0.04<br>p > 0.05 for all other sleep and delirium analyses |
| Simeone, 2018, Italy <sup>46</sup> | Observational, correlational | SICU (cardiac)<br>n=89<br>All patients aged $\geq 18$ years who underwent cardiac surgery with ICU stay longer than 24 hours, excluding history of psychologic disease or psychogenic drug use, visual disturbances, hearing disorder, RASS $\leq 4$ | Exposure to natural sunlight<br>Presence of a sleep disorder | Delirium: RASS followed by CAM-ICU<br>Light: whether patient was exposed to natural sunlight<br>Sleep: whether patient has pre-existing sleep disorder | Significantly more patients with delirium were not in a location with sunlight<br>Significantly more patients with delirium had a sleep disorder | Light, univariate<br>$X^2 = 9.737$<br>p = 0.32<br>Light, multivariate<br>RRR 0.367<br>95% CI (0.090-1.494) p = 0.034<br>Sleep, univariate<br>$X^2 = 13.934$<br>p < 0.001<br>Sleep, multivariate<br>RRR 5.493<br>95% CI (1.255-24.047) p = 0.024 |

**Abbreviations:** AASM: American Academy of Sleep Medicine; CABG: coronary artery bypass graft; CAM-ICU: Confusion Assessment Method for the Intensive Care Unit; CBO: clinical behavioral observation; CI: confidence interval; CNS: central nervous system; dB: decibel; df: degrees of freedom; DSM-III: Diagnostic and Statistical Manual of Mental Disorders, 3rd Edition; DSM-III-R: Diagnostic and Statistical Manual of Mental Disorders, 3rd Edition, Revised; DSM-IV: Diagnostic and Statistical Manual of Mental Disorders, 4th Edition; ECG: electrocardiography; EEG: electroencephalography; EMG: electromyography; EOG: electrooculography; GCS: Glasgow Coma Scale; HCU: high intensive care unit; ICD-9-CM: International Classification of Diseases, Ninth Revision, Clinical Modification; ICDSC: Intensive Care Delirium Screening Checklist; ICU: intensive care unit; Leq: equivalent continuous sound pressure level; MICU: medical intensive care unit; MMSE: Mini-Mental State Examination; MV: mechanical ventilation; NEECHAM: Neelon and Champagne Confusion Scale; NIV: non-invasive ventilation; OR: odds ratio; PSG: polysomnography; RASS: Richmond Agitation and Sedation Scale; RCSQ: Richards-Campbell Sleep Questionnaire; REM: rapid eye movement; RRR: relative risk ratio; SAS: Riker Sedation-Agitation Scale; SICU: surgical intensive care unit; $X^2$ : chi-squared test.

**Table 2.** Summary of Characteristics and Findings of Reviewed Interventional Studies.

| Study<br>(Author,<br>Year,<br>Country) | Study<br>Purpose | Study<br>Design | Study Setting<br>Population<br>Subjects Characteristics | Intervention Details | Outcomes<sup>1</sup> (Methods of<br>Assessment) | Findings |
| --- | --- | --- | --- | --- | --- | --- |
| **Architectural design modification** |  |  |  |  |  |  |
| Johansson,<br>2018,<br>Sweden <sup>18</sup> | To assess<br>feasibility and<br>effect of a<br>modified ICU<br>room on noise<br>and delirium | Quasi RCT<br>(feasibility<br>study) | ICU (General)<br>n=31: 25 control, 6<br>intervention<br>Adult, ICU stay ≥48 hrs | Modified ICU room to<br>control noise: Installed drop<br>ceilings with low frequency<br>noise absorber, plain room<br>design, kept mobile medical<br>equipment in room only if<br>required<br>Control group: same ICU<br>room with no modification | Delirium prevalence(CAM-ICU)<br>Level of noise (Microphone<br>located 10cm below ceiling,<br>and 130-160 cm from wall,<br>attached to a sound-card,<br>recorded 30s intervals of A, C,<br>and Z weighted noise levels) | Reported study as feasible<br>with required improvement in<br>randomization, noise<br>measurement process, and<br>delirium assessment<br>No statistical analysis<br>performed due to small sample<br>size; however intervention<br>resulted in slight lower<br>reverberation time and higher<br>speech clarity |
| Zaal, 2013,<br>Netherlands<br><sup>47</sup> | To explore<br>effect of ICU<br>environment<br>on incidence,<br>and course of<br>delirium | Pre- post<br>Intervention | ICU (Mixed)<br>n=130: 55 control, 75<br>intervention<br>Adult, ICU stay ≥24 hrs,<br>excluded unresponsive<br>patients (RASS <-3 or GCS<br>≤ 8) in ICU | Single-bed room ICU with<br>more daylight and less<br>noise exposure: Use of<br>sound absorbers, glass<br>sliding doors, optimized<br>alarm system sending<br>filtered alarms to staff cell<br>phones, remotely controlled<br>monitors, sufficient daylight<br>with view, warm colored<br>room design<br>Control group: multi-bed<br>room, beds separated by<br>curtains | Delirium incidence, and<br>duration (CAM-ICU)<br>Delirium severity (non-validated DSI)<br>Level of light (Light-sensor<br>placed 1m from bed's head,<br>recorded 30s intervals of light<br>intensity in Volts) | No significant effect on<br>incidence of delirium<br>Decreased number of days<br>with delirium<br>No effect on severity of<br>delirium<br>Increased daylight exposure,<br>but no effect on night-time<br>light exposure |
| **Noise modification interventions** |  |  |  |  |  |  |
| van de Pol,<br>2017,<br>Netherlands<br><sup>14</sup> | To test effect<br>of nocturnal<br>sound-reduction<br>protocol on<br>delirium<br>incidence and<br>sleep quality in<br>ICU | Pre- post<br>Intervention<br>(interrupted<br>time series) | ICU (Medical and Surgical)<br>n=421: 211 control, 210<br>intervention<br>Adult, non-delirious,<br>RASS<-3 for > 50% of ICU<br>stay, ICU stay≥24 hrs | Nocturnal sound-reduction<br>protocol: Lowered staff and<br>devices noise, clustered<br>care-activities, closed doors<br>and earplugs in non-delirious patients, limited<br>and clustered care activities<br>Control group: usual care | Delirium incidence (ICDSC)<br>Sleep (RCSQ)<br>Level of noise (Perceived noise<br>item of RCSQ, and Sound<br>meter with microphone located<br>near bed's head, recorded 1s | Decreased delirium incidence<br>No improvement in sleep<br>quality<br>Reduced perceived night-time<br>noise. Noise pressure levels<br>not compared between the two |
|  |  |  |  |  | intervals of A weighted noise levels) | groups due to unusable pre-intervention values<br>Reduced use of sleep medication, no effect on delirium medication |
| Van Rompaey, 2012, Belgium <sup>48</sup> | To evaluate effect of sleeping with earplugs on prevention of delirium in ICU | RCT | ICU (Medical, surgical, cardiosurgical)<br>n=136: 67 control, 69 intervention<br>Adult, non-delirious, non-sedated, non-intubated, GCS≥10, no dementia, ICU stay >24hrs | Earplugs during sleep from 2200 to 0600<br>Control group: No earplugs | Delirium incidence (NEECHAM)<br>Sleep perception (Non-validated simplified sleep questionnaire with five dichotomous questions) | No effect on incidence of delirium, but reduced risk of confusion, and increased time to cognitive disturbance onset<br>Improved sleep quality |
| **Light modification interventions** |  |  |  |  |  |  |
| Estrup, 2018, Denmark <sup>15</sup> | To explore delirium incidence, and risk factor and effect of circadian light on delirium in ICU | Retrospective cohort study | ICU (Mixed medical and surgical)<br>n=183<br>Adult, non-sedated patients with available CAM-ICU scores, without any coma: RASS of -5 or -4, or severe dementia | Circadian Lighting: Supplemental lighting system delivered light with strongest intensity of 4000lx and most blue component from 0700 to 1200, and minimum of 50lx at 2030<br>Control group: Regular lighting | Delirium Incidence (CAM-ICU, and use of Haloperidole) | No effect on incidence of delirium<br>Reported age and dexedetomidine as risk factors for delirium |
| Pustjens, 2019, Netherlands <sup>49</sup> | To test effect of dynamic lighting on delirium and | Retrospective cohort study | CCU<br>n=748: 379 control, 369 intervention<br>Adult non-sedated patients with ICU stay of 24 hrs or more | Circadian Lighting: Ceiling mounted LED panels delivered light with color temperature between 2700 and 6500 K at varying intensities (peak 750lx)<br>Control group: Regular lighting | Delirium incidence (DOSS, and CAM) | No effect on incidence of delirium |
| Simons, 2016, Netherlands <sup>50</sup> | To assess effect of dynamic lighting on incidence and duration of delirium | RCT | ICU (Mixed medical & surgical)<br>(n=714: 360 control, 354 intervention)<br>Adult, ICU stay>24 hrs, both intubated and non-intubated without impairments preventing | Artificial high-intensity dynamic lighting application: Ceiling mounted fluorescent tubes delivered light with alteration in color temperature and intensity: blueish-white light up to 4300 K, and 1700 lx from 0900 to 1600, except | Delirium incidence, and duration (CAM-ICU)<br>Level of light (Photometer placed at 2m height on wall near bed's head, recorded 15min intervals of light intensity in lx) | No significant effect on incidence of delirium, or number of delirium-free days<br>Increased mean cumulative daytime lighting |
|  |  |  | delirium assessments were included | intensity of 300 lx from 1130 to 1330<br>Control group: Usual lighting: 300 lx, 3000 K |  |  |
| Taguchi, 2007, Japan<sup>51</sup> | To evaluate effect of BLT on post-operative circadian optimization and delirium | RCT (Pilot) | ICU (surgical)<br>n=11: 5 control, 6 intervention<br>Middle-aged, or aged post-operative esophageal cancer patients, with no mental disorders were randomized after extubation | BLT: 5000lx from 0730 to 0930 for 3 of the post-operative days, by a self-stand or a table-top illuminator<br>Control group: Usual lighting: 600-1000lx | Delirium incidence (non-validated Japanese NEECHAM)<br>Sleep/Circadian rhythm (Activity levels and rhythm recorded by ankle accelerometers and memory heart rate recorder) | Decreased post-operative delirium rate on day 3 of the BLT, but no overall significant effect on delirium incidence<br>Non-significant decrease in activity level during sleep |
| Ono, 2011, Japan<sup>52</sup> | To evaluate effect of BLT on post-operative circadian optimization and delirium | RCT (Pilot) | ICU (surgical)<br>n=22: 12 control, 10 intervention<br>Adult post-esophagectomy patients, who anticipated to be extubated the day after surgery, were randomized after extubation | BLT: 2500 to 5000lx from 0730 to 0930 for 4 days (0730-0745: 2500lx, 0745-0800: 4000lx, 0800-0900: 5000lx, 0900-0915: 4000lx, 0915-0930 2500lx), by a self-standing L-shaped illuminator to maintain lighting in front of patient's face<br>Control group: usual lighting | Delirium incidence (Validated Japanese NEECHAM)<br>Sleep/Circadian rhythm (Activity levels and rhythm recorded by ankle accelerometers and memory heart rate recorder ) | Non-significant lower rate of post-operative delirium<br>Decreased amount of activity during sleep on the nights of days 4 and 5 |
| Potharajaroen, 2018, Thailand<sup>55</sup> | To evaluate effect of BLT on post-operative delirium | RCT | ICU (surgical)<br>n=62: 31 control, 31 interventions<br>Adult patients ≥ 50 years, with an APACHE II Score ≥ 8 with no current coma or delirium, a life-time history or current diagnosis of delirium, neuro-degenerative , neuroinflammatory, or psychiatric disease. | BLT: Bright Light therapy, 5000lx from 0900 to 1100 for 3 days, at a distance of 1.40 m from the patient's face<br>Control group: usual lighting: 500lx | Delirium incidence (CAM-ICU)<br>Sleep (Assessing insomnia by ISI) | Decreased delirium incidence<br>Higher ISI score was associated with development of delirium, and BLT lowered ISI scores. |
| Yang, 2012, South Korea<sup>53</sup> | To determine impact of BLT with antipsychotic | Randomized open parallel group | Consulting psychiatry division of a general hospital | BLT as adjunctive treatment with risperidone: 10,000 lx from 0700 to 0800 for 5 | Delirium severity (DRS, MDAS) | Decreased delirium severity<br>Improve total sleep time and sleep efficiency |
| | treatment in delirious patients | | n=36 (16 control, 20 intervention)<br>Referred hospitalized adults to psychiatry division of a general hospital with DRS≥ 12 without any other axis I disorders on DSM-IV or history of antipsychotics or benzodiazepines use before screening | days by a height-adjustable light box<br>Control group: Risperidone | Sleep (Sleep log with total sleep time, efficiency, onset latency, awake times questions) | |
| Chong, 2013, Singapore <sup>54</sup> | To examine whether the GMU program improved sleep, cognitive, and functional outcomes in delirious patients. | Prospective cohort study | GMU<br>n=228<br>Adult delirious patients >65 years old, without coma, or terminal illness, or contraindications to BLT (manic disorders, severe eye disorders, photosensitive skin disorders, or photosensitizing use) | BLT: 2000- 3000lx from 1800 to 2200 delivered by ceiling lights in addition to HELP protocol | Delirium severity (CAM, DRS-98, locally validated CMMSE)<br>Sleep (Sleep log with total sleep time, number of awakenings, number and length of sleep bouts questions) | No significant effect on DRS severity scores, but improved DRS sleep-wake disturbance sub-score, No significant improvement on CMMSE scores<br>Improved mean total sleep time, and functional status score during management of delirium |
| **Environmental modification targeting both noise and light** |  |  |  |  |  |  |
| Demoule, 2017, France <sup>56</sup> | To evaluate effect of earplugs and eye mask on sleep in ICU | RCT | ICU (General)<br>n=43; 28 control, 15 intervention<br>Adult, non-sedated, Ramsay Sedation Scale <3, no history of sleep disorder, neurological impairment, encephalopathy, sepsis, ICU stay >48hr | Earplugs and eye-masks during sleep from 2200 to 0800<br>Control group: No earplugs or eye mask | Delirium incidence (CAM-ICU)<br>Sleep (PSG on first day of study, Self-reported sleep quality by simplified visual analogue scale (VAS; 10 cm horizontally) at discharge, and by Pittsburgh Sleep Quality Index at day 90) | No effect on delirium<br>No effect on sleep proportion of N3, but improved sleep quality only by increasing duration of N3 stage and reducing long awakenings in compliant subjects. |
| McAndrew, 2016, USA <sup>57</sup> | To evaluate effect of quiet time on delirium, sedation level, and physiologic measures in mechanically ventilated patients | Prospective cohort study | ICU (Medical)<br>N=72<br>Mechanically ventilated adults until extubated | Quiet time from 1400 to 1600; Dimmed lights, closed window shades, TVs off, closed doors, clustered care-activities | Presence of delirium (CAM-ICU)<br>Sleep (Nurse perception of patient's sleep by an investigator created tool with uninterrupted sleep time, and overall quality of sleep questions) | No significant effect on delirium; however reported no increase in delirium<br>Improved sleep perception moderately<br>Improved respiratory rates, and nursing satisfaction of quiet time protocol |
| Patel, 2014, UK <sup>7</sup> | To test a non-pharmacologic bundle with environmental noise and light reduction components on delirium and sleep | Pre- post Intervention | ICU (Medical & surgical)<br>N=338; 167 control, 171 intervention<br>Non-delirious, non-sedated adults with $\geq 1$ ICU night, and no sleep, or cognitive, or neurologic disorder | Multidisciplinary intervention from 2300 to 0700; Limited bedside conversation, clustered care-activities, minimized devices noise levels, dimmed lights, earplugs and eye mask, patient orientation, early mobilization, and sedation targets.<br>Control group: usual care | Delirium incidence and duration (CAM-ICU)<br>Sleep quality (RCSQ, and the Sleep in Intensive Care Questionnaire)<br>Level of noise<br>Level of light<br>(Two environmental meters placed centrally; mean level of noise reported in dB, illuminance reported in lx, no more information available) | Decreased delirium incidence and duration<br>Increased sleep quality, decrease daytime sleepiness<br>Decreased level noise<br>Decrease level of light |
| Kamdar, 2013, USA <sup>58</sup> | To determine impact of a multi-faceted quality improvement program on ICU delirium, and sleep | Pre- post Intervention | ICU (Medical)<br>n=300; 122 control, 178 intervention<br>Adult patients with $\geq 1$ ICU night, and discharge to an inpatient ward bed or pending discharge directly from the ICU, without $\geq 1$ night in another ICU during the admission, any cognitive disorder, or sustained alcohol or drug abuse, cardiac arrest during admission, any other ICU discharge $> 96$ hours prior to assessment | Multi-faceted sleeping promotion protocol; 3 additive stages of 1) quiet time, and realignment of circadian rhythm, 2) earplugs, eye-masks, and soothing music, 3) pharmacological targets to reduce sedatives.<br>Control group: usual care | Delirium incidence (CAM-ICU)<br>Sleep (RCSQ) | Decreased incidence of delirium<br>No effect on quality of sleep ratings |

**Abbreviations:** ICU: Intensive care unit; RCT: Randomized control trial; hr: Hours; CAM-ICU: Confusion Assessment Method for the ICU; LAeq: A-weighted equivalent continuous sound level, LAFMin: A weighted, Fast time (125ms) weighted minimum sound Level, LAFMax: A weighted, Fast time (125ms) weighted maximum sound Level, LCeq: C-weighted equivalent continuous sound level, LCPeak: C-weighted peak sound level, LZPeak: Peak sound level without frequency weighting, T20: Reverberation time, C50: Clarity, RASS: Richmond Agitation and Sedation Scale, GCS: Glasgow Coma Scale/Score, DSI: delirium severity index, ICDSC: Intensive Care Delirium Screening Checklist, RCSQ: Richards-Campbell Sleep Questionnaire, dBA: A-weighted decibels, NEECHAM: Neelon and Champagne Confusion Scale, CCU: Coronary care unit, LED: Light-emitting diode, DOSS: Dutch version of the Delirium Observation Screening, CAM: Confusion Assessment Method, lx: Lux, K: Kelvin, min: minutes, BLT: Bright light therapy, ISI: Insomnia severity score, DRS: Delirium Rating Scale, DSM-IV: on Diagnostic and Statistical Manual of Mental Disorders, 4<sup>th</sup> edition, MDAS: Memorial Delirium
Assessment Scale, GMU: Geriatric Monitoring Unit (A specialized delirium management unit), HELP: Hospital Elder Life Program (standardized protocols to manage cognitive impairment, sleep deprivation, immobility, visual impairment, hearing impairment, and dehydration), DRS-98: Delirium rating scale-R98, CMMSE: Chinese Mini–Mental State Examination, PSG: Polysomnography,
<sup>1</sup> Outcomes of interest including delirium related outcomes, sleep quality, sound pressure levels, and light intensity levels, has listed in this table.
<sup>2</sup> Details of measured noise and light, such as devices, location, and frequency has not been discussed in detail in this table.

### Effect of environmental risk factors on delirium

Of the 21 observational studies, two analyzed for association between delirium and noise ^19, 20^, five for light factors and delirium ^13, 29–32^, 12 for sleep and delirium ^33–44^, and two evaluated multiple factors (noise, light, and/or sleep with delirium) ^45, 46^. Study population ranged in size from 7 to 6660 participants, and the vast majority of studies were done in an ICU (17 of 21 studies) ^13, 19, 30–33, 35–39, 41, 42, 44–46^. The remaining four studies did not specify a ward and were performed in a general hospital setting ^29, 34, 40, 43^. Study details and reported statistical results are in Table 1.

#### Noise

Although ICU noise is a suggested predictor for delirium development, two of the three investigating studies found no significant association between ICU noise levels and delirium development ^19, 20^. One study assessed A-weighted sound levels with subjective patient reports on ICU noise ^20^. They found no correlation between A-weighted equivalent continuous (LAeq) or maximum (LAmax) noise pressure levels and delirium, while patients’ responses about ICU sounds spread evenly over a spectrum from scary to non-disturbing ^20^. In comparison, Knauert et al. ^19^ evaluated equivalent continuous sound pressure level (Leq) and peak sound occurrences for both A-weighted and C-weighted measurements, finding no correlation with delirium development. There are no industry-standard recommendations for C-weighted levels, but LAeq and LAmax values from both studies were higher than recommended by the WHO ^17, 19, 20^. In contrast, a study by Davoudi et al. ^45^ found average night-time sound pressure levels were significantly higher for patients with delirium ^45^. However, they did not provide exact decibel measurements to compare with recommended WHO levels, likely because they were reporting preliminary findings for a larger cohort study unpublished at the time of this review ^45^.

#### Light

Abnormal lighting cycles are another suggested contributor to delirium ^59^. Seven of the reviewed studies considered exposure to natural sunlight and any statistical relationships with delirium ^13, 29–32, 45, 46^. There were two approaches to analysis: effects of windows on delirium incidence ^13, 30, 32, 45, 46^ and association with admission season ^45, 46^. Findings were mixed across the studies, suggesting no easily provable relationship between natural light exposure and delirium occurrence. Two window and one seasonal study found no statistical association between delirium and windows or season of admission/duration of preadmission sunlight exposure, respectively ^30–32^. Kohn et al. ^30^ compared windowed versus non-windowed rooms in the medical ICU, and natural versus industrial window views in the surgical ICU. ^30^. They also investigated impact of half-sized versus full-sized windows, finding no association between delirium incidence and any of these factors ^30^. Similarly, Smonig et al. found no difference in delirium incidence between patients admitted to windowed versus non-windowed rooms while proving windowed rooms retained natural circadian light variations and non-windowed rooms did not ^32^. In the seasonal study, Simons et al. investigated effect of admission season with delirium and found no correlation ^31^. A simultaneous assessment found no correlation between preadmission cumulative sunlight exposure and delirium incidence for three photoperiods (7, 28, and 60 days pre hospital admission) ^31^.

In comparison to studies showing no association between natural sunlight exposure and delirium occurrence, three window studies and one seasonal study found a significant correlation ^13, 29, 45, 46^. In the window studies, Simeone et al. ^46^ associated lack of natural sunlight with delirium while Van Rompaey et al. ^13^ found absence of visible daylight led to higher risk of delirium. Davoudi et al. ^45^ examined pervasive sensing of ICU patients, finding measured light intensity in windowed rooms was significantly different between patients with and without delirium ^45^. Additionally, a study on seasonal impact on delirium diagnosis by Balan et al. found a higher incidence of delirium among patients admitted in winter compared to summer ^29^.

#### Sleep

Disrupted sleep-wake cycles are associated with altered mental state in hospitalized patients, and are connected with delirium ^60^. In this review, 14 studies ^19, 33–37, 39–46^ assessed sleep and delirium with two main methodologies: objective measurements of physiological sleep phases and subjective reports by staff or patient. Five studies objectively measured sleep quality using overnight polysomnography (PSG) or a Zeo wireless sleep monitor ^19, 33, 35, 41, 42^, while eight assessed staff reports of behavioral observations and/or self-reports by patients ^33, 34, 36, 37, 40, 44–46^. One study compared both methods [33], and two did not specify their method of measurement, only stating that they evaluated the relationship between sleep deprivation and delirium ^39, 43^.

Similar to the articles on natural light exposure, association studies for sleep and delirium have mixed findings, but lean towards disrupted sleep being a delirium predictor. Six of 14 studies found no relationship between sleep and delirium: two PSG studies ^19, 33^, three using subjective measures ^33^, and one with unspecified method ^36, 39, 45^. One study found no difference in rate of delirium between patients with typical and atypical sleep on PSG ^38^, while another by Boesen et al. also found no difference in atypical PSG results between patients who did or did not develop delirium ^33^. They compared PSG results with clinical behavioral observations and were only able to ascertain that the more pathological the patient and electroencephalogram findings, the less association with observed sleep ^33^. A study using the Richards-Campbell Sleep Questionnaire (RCSQ) found no significant correlation between perceived sleep quality and delirium, nor any significant relationship when asking how disruptive noise was to sleep ^36^. The study by Davoudi et al. ^45^ used the Freedman Sleep Questionnaire and found no correlation between overall sleep quality and delirium, although they noted patients with delirium were more likely to have difficulty falling asleep and find night-time lighting disruptive ^45^. The last study did not detail their methodology, but found delirium was not significantly related to sleep deprivation ^39^.

Of nine studies showing statistical correlation between sleep and delirium, three used electronic sleep monitoring ^35, 41, 42^, five subjective survey measures ^34, 40, 44, 46, 61^ and one did not specify methodology ^43^. One study found atypical sleep on PSG was significantly tied to increased delirium, while another PSG study found delirium was associated with severe REM reduction ^35, 41^. A third study used a novel sleep monitoring device and found a relationship between lack of rapid eye movement (REM) sleep and delirium ^42^. Their results must be taken in the context of the device being commercially unavailable (Zeo wireless sleep monitor), and the authors not reporting statistical analyses. Among remaining positive correlational studies, two had patients self-report sleep satisfaction and quality and both saw significantly poorer responses when comparing patients who developed delirium with those who did not ^34, 44^ Two studies used nursing staff observing clinical behaviors and found sleep disturbances were positively linked to higher likelihood of developing delirium ^37, 40^. Two studies found an association between delirium incidence and sleep deprivation (methodology not specified) ^43^, and between sleeping disorders and delirium development ^46^.

### Effect of environmental interventions on delirium prevention and treatment

Sixteen studies evaluated effects of a modified environment on delirium prevention or management ^7, 14, 15, 18, 47–58^ (Table 2). Half were randomized control trials (RCT)^18, 48, 50–53, 55, 56^, while half used different study designs including: before-after ^7, 14, 47, 58^, retrospective cohort ^15, 49^, and prospective cohort ^54, 57^. Sample sizes varied from 11 to 748. Interventions focused on controlling environmental risk factors, including noise and light exposure, disrupted circadian rhythm, and sleep (Figure 2). We categorized these interventions into four modification types: architectural design ^18, 47^, environmental noise ^14, 48^, environmental light ^15, 49–56^, and environmental modification bundles with noise and light components ^7 56–58^. A summary of environmental interventions on delirium and reported statistical results are presented in Table 3.

**Figure2.**
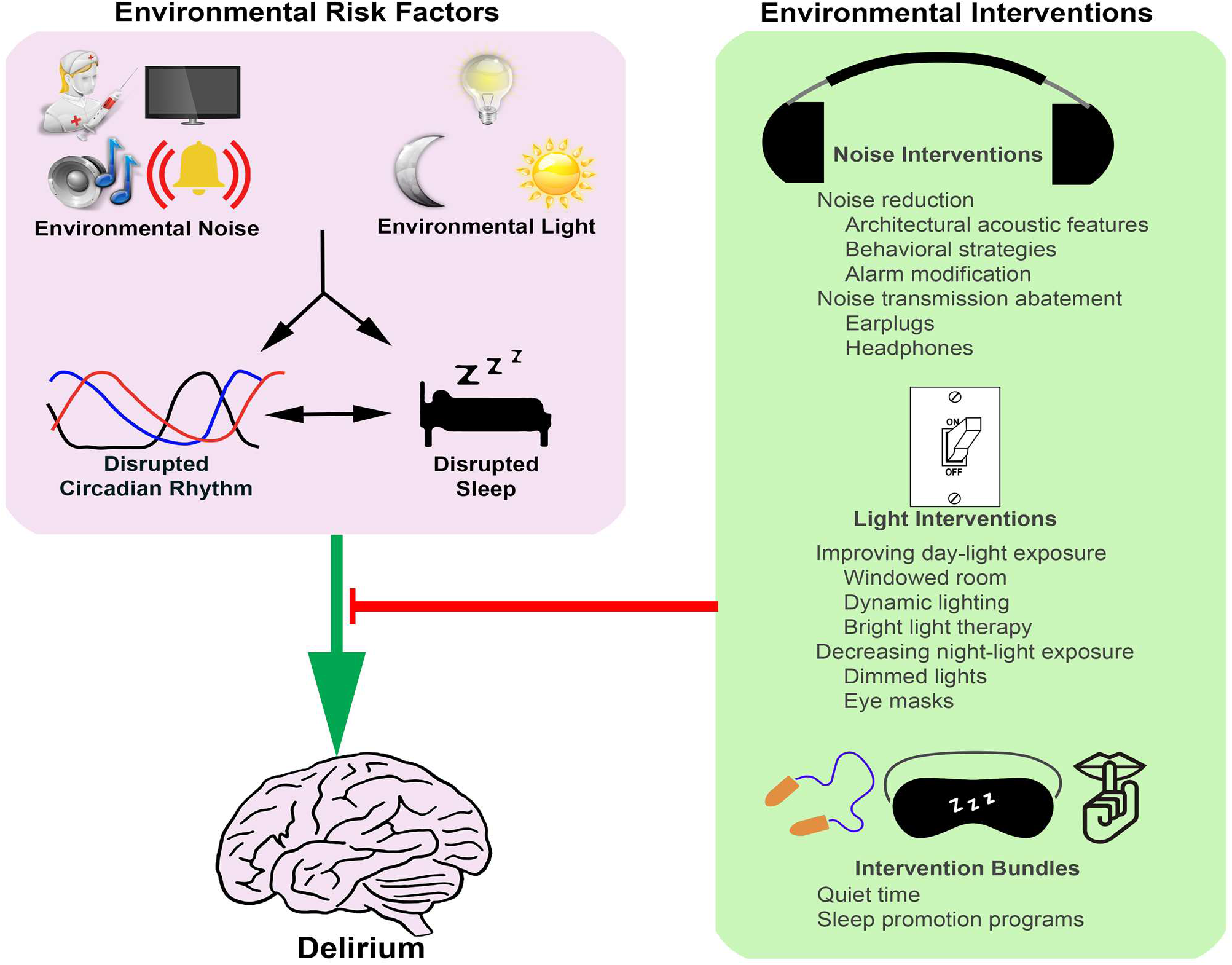
Environmental Risk Factors for Delirium, and the Mitigation Strategies.

**Table 3.**
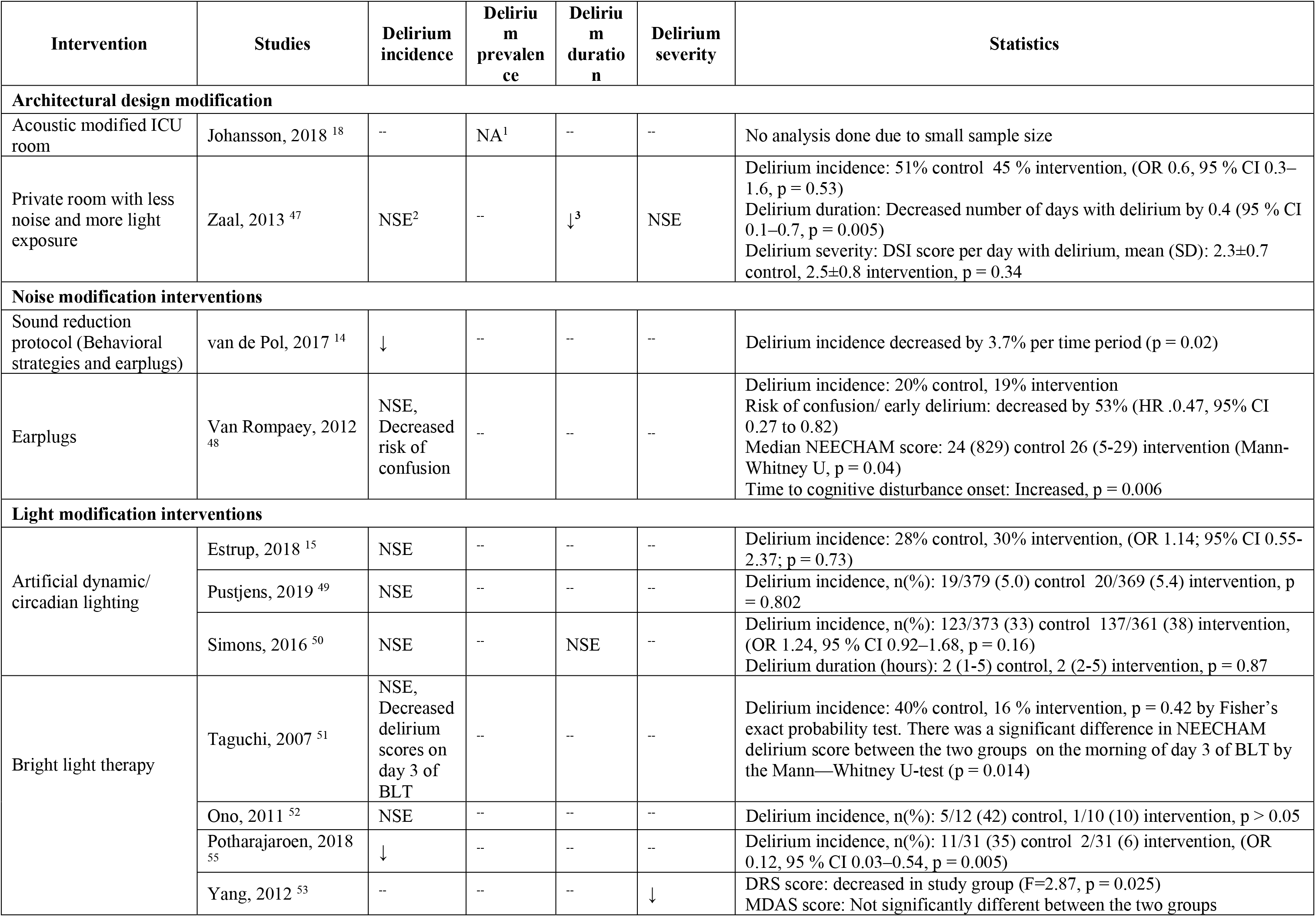
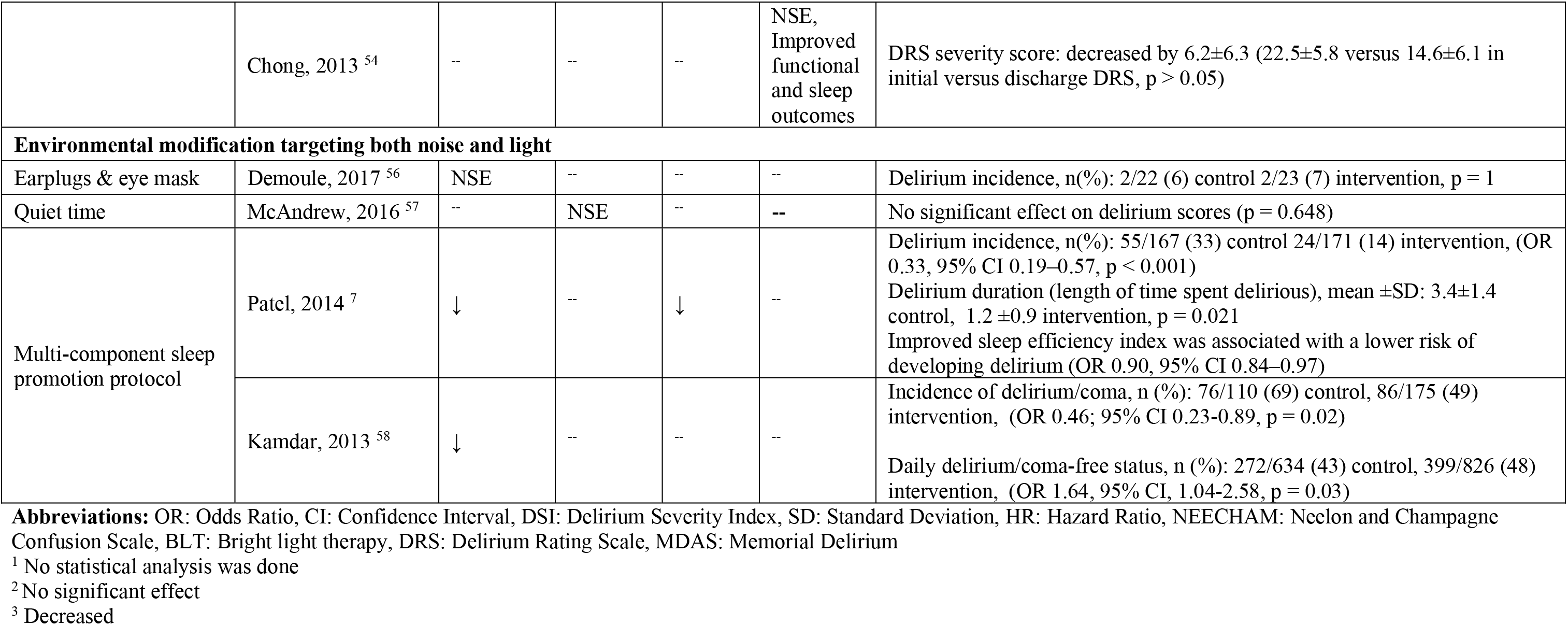
Effectiveness of Environmental Interventions on Delirium.

#### Architectural design

In this review, two of the studies [18, 47] explored a modified ICU design. One study altered the acoustical design of the ICU ^18^, whereas the other used a multi-aspect architectural design intervention ^47^. Results were mixed, but subtly suggest the benefit of architectural designs that consider acoustic features. Zaal et al. ^47^ assessed patient outcomes in a multi-bed ICU room with less natural light and more noise exposure versus a private room with improved daylight and reduced noise by sound absorbers, glass sliding doors, optimized alarms, and remotely controlled monitors. There was no effect on delirium incidence or severity, but they found a reduction of delirious days in the study group by 0.4 (95% confidence interval (CI) 0.1–0.7, p = 0.005). Another quasi-randomized study ^18^ conducted noise reduction by refurbishing an ICU room. They installed a wall-to-wall drop ceiling, low frequency sound absorbers, and used a visually plain design. The study deemed feasible, requiring improvements in noise measurements and delirium assessments. Given the small sample size (n=31) and feasibility nature of study, no further statistical analysis of outcomes was performed; Delirium developed in 33% (2/6) versus 25% (5/25) of study versus control patients. There was a slight reduction in noise reverberation and increase in speech clarity in the modified room, though sound levels remained higher than the WHO recommendations ^17^.

#### Noise modification

In this review, there were two approaches to mitigate patient exposure to excessive sound. One was to reduce source noise by utilizing behavioral strategies and device/alarm optimization. The other was noise abatement by earplugs. No studies investigated impacts of behavioral modification on delirium as an independent intervention, but this strategy was used as part of an environmental modification bundle in 4 studies ^7, 14, 57, 58^. Earplugs were mostly a component of an environmental bundle ^7, 14, 56, 57^, though one study evaluated the effect of earplugs as a single-component intervention ^48^. One article implemented a combination of behavioral strategies and earplugs to reduce excessive noise ^14^ There were mixed findings across studies with noise modification component(s), but results suggest behavioral strategies and earplugs together might help delirium prevention, particularly as part of a multi-disciplinary program targeting environmental risk factors. However, implementation of sustained behavioral changes and tolerability of earplugs remain challenges ^56^.

Van de Pol et al. ^14^ analyzed the impact of noise reduction on 421 non-delirious ICU patients in an interrupted time series before-after study. They used earplugs and behavioral strategies, including limited bedside conversations, lowered voices, grouped care activities, optimized alarm settings, minimized alarm volume, and closed room doors. Reported noise levels were still higher than the WHO limit post-intervention ^17^, however there was a significant decrease in delirium incidence by 3.7% per time interval (p = 0.02), and reduction in sleep medication usage (p < 0.0001) in the study group. Perceived night-time noise was improved, but with no effect on sleep quality or use of delirium medication. Van Rompaey et al. show associations between environmental noise, sleep perception, and delirium ^48^. They conducted a randomized control trial on 136 non-delirious ICU patients and found use of earplugs (from 2200 to 0600) reduced risk of confusion or delirium by 53% (hazard ratio 0.47, 95% CI 0.27-0.82) and improved sleep perception.

Our full-text review and data extraction appraised articles studying single-component noise control strategies, such as behavioral programs ^62–66^, earplugs or noise cancelling headphones ^67–71^, and headphones equipped with an alarm filtering system ^72^; however these were not included since they reviewed the impact of interventions on the level of noise or quality of sleep, but delirium was not reported as an outcome (Excluded studies; Supplementary Table 3).

#### Light modification

Light interventions were implemented in an attempt to realign circadian rhythms by reducing night-time exposure and/or improving natural or artificial daylight exposure.

##### Reduction of nocturnal light exposure

In this review, eye mask use ^7, 56, 58^, and overnight light dimming ^7, 57, 58^ were encouraged as part of an environmental modification bundle to reduce night-time light exposure. No studies evaluated effects of less nocturnal light exposure on delirium as single interventions.

##### Improving natural daylight exposure

Three observational studies ^30, 32, 73^ and one before-after study ^47^ investigated improved natural lighting via windows. They compared patient outcomes in rooms with a window or larger-sized windows versus windowless or smaller-sized windows, respectively. No observational studies suggested association between improved natural lighting and delirium ^30, 32, 73^. Zaal et al. ^47^ demonstrated reduction in delirium duration, comparing patients in private rooms with more natural light versus less bright multi bed rooms; however, there was no difference in delirium incidence or severity between groups.

##### Improving artificial daylight exposure

Eight studies examined effect of improved daylight exposure via artificial lighting, of which three used an artificial circadian lighting system ^15, 49, 50^, and five used bright light therapy (BLT) ^51–55^. None of the three studies implementing artificial dynamic or circadian lighting revealed significant effects on delirium. BLT studies had mixed results; three studies significantly improved delirium prevention or management, while other two showed a non-significant tendency to reduce delirium rates.

A retrospective cohort study of 183 non-sedated ICU patients by Estrup et al. ^15^ used a circadian lighting system from 0700 to 2300 which varied in intensity and color temperature. During the morning, light intensity was greatest, up to 4000 lux (lx), and the amount of blue light strongest. As the day progressed, light intensity decreased and color temperature shifted towards warmer tones until no blue light was present. There was no improvement in delirium incidence, and no association between receiving circadian lighting and delirium incidence (odds ratio (OR) 1.14; 95% CI 0.55, 2.37; p = 0.73). Pustjens et al. ^49^ retrospectively studied a cohort of 748 non-sedated patients. They implemented a dynamic lighting system consisting of two ceiling-mounted light-emitting diode (LED) panels which delivered variable intensities of light (peak of 750 lx) with a color temperature between 2700 and 6500 Kelvin (K). There was no effect on delirium incidence. Another RCT by Simons et al. ^50^ measured effects of a dynamic lighting application (DLA) in 734 ICU patients. DLA was administered through ceiling-mounted fluorescent lights which delivered a variety of bluish-white light from 0700 to 2230 with a maximum intensity of 1700 lx and a maximum temperature of 4300 K between 0900 and 1600, except between 1130 and 1330 when light intensity was 300 lx. This study was terminated before reporting final results, but preliminary analysis demonstrated delirium incidence of 38% versus 33% in control versus study patients, with no significant improvement on delirium incidence or duration in the study group.

Four studies investigated use of BLT as a single-component intervention to prevent ^51, 52, 55^ or treat ^53^ delirium, while one study used BLT as an element of a multi-component bundle to manage delirium ^54^. BLT consisted of exposure to high intensity light (2000 to 10000 lx) for one to four hours daily. Three studies used a peak intensity of 5000 lx ^51, 52, 55^. Taguchi et al. ^51^ conducted a randomization pilot study on 11 post-operative patients, utilizing a daily light intensity of 5000 lx from 0730 to 0930 for days 2 through 5 post-surgery. Delirium assessment scores decreased on day 3 of BLT (p = 0.014), but there was no significant effect to overall delirium incidence (16% versus 40% study versus control group, p = 0.42). In another RCT, Ono et al. ^52^ applied BLT on 22 post-operative patients, for two hours from 0730 to 0930 for four days. Light intensity started at 2500 lx, increasing to 5000 lx, then decreasing to 2500 lx. There was a non-significant tendency towards lower rates of delirium in the study group (1 of 10 patients) versus control group (5 of 12 patients), while BLT significantly reduced the amount of activity during sleep of day 4 and 5. Potharajaroen et al. ^55^ studied 62 post-operative patients by implementing BLT at 5000 lx with a constant intensity from 0900 to 1100. Eleven of 31 control patients versus 2 of 31 patients in the intervention group developed delirium. There was a significant association between BLT and decreased delirium incidence (OR 0.12, 95% CI 0.03–0.54, p = 0.005). A study by Yang et al. ^53^ on 36 delirious patients used a higher light intensity (10000 lx) over a shorter period (0700 to 0800). This study investigated the use of BLT as an adjunctive treatment of delirium with risperidone. They found a significant decrease in delirium severity in patients receiving BLT in addition to risperidone (DRS 23.9 ± 4.9 versus 20.6 ± 3.6 in control versus study group, p = 0.03). Chong et al. ^54^ studied 228 delirious elderly patients admitted to a delirium management unit. They incorporated lower intensity BLT as part of their multi-component program, and exposed patients to 2000 to 3000 lx of light for four hours from 1800 to 2200 daily. They reported significant improvement in total sleep time and functional outcomes during treatment of delirious patients.

#### Intervention bundles (combination of light and noise modification)

##### Earplugs and eye mask

One reviewed study explored effects of earplugs and an eye mask on delirium ^56^, while two others used earplugs and an eye mask as part of their interventional bundle ^7, 58^. All three decreased incidence of delirium, but had different effects on sleep quality. Demoule et al. ^56^ conducted a RCT on 43 non-sedated ICU patients to investigate impact of sleeping with earplugs and an eye mask from 2200 to 0800 on patient outcomes. They found no improvement in delirium incidence or duration or architecture of sleep in the study group, regardless of patient compliance using the equipment. Although compliant study subjects experienced improved sleep with longer N3 (deeper sleep) duration and a lower number of prolonged awakenings, there was no significant change in delirium incidence. There were several articles in our initial screening reporting improved perceived noise or sleep quality with use of earplugs and eye mask, however those were excluded since none reported results on delirium ^74–77^ (Excluded studies; Supplementary Table 3).

##### Quiet time, and sleep promotion bundles

Quiet time is a specific amount of time during which modifiable noise and light is actively reduced. Our review included three studies installing quiet time as the single interventional element ^57^ or as a part of a sleep promotion bundle ^7, 58^. Core elements of quiet time were behavioral strategies, minimized bedside activity by clustering care, reduced volume of devices/alarms, and dimmed lights ^7, 57, 58^. The study that implemented daytime quiet time failed to show significant effects on delirium ^57^, while two sleep promotion studies decreased delirium incidence using nocturnal quiet time combined with components such as earplugs, eye masks, and pharmacological targets ^7, 58^. Although the multi-component sleep promotion trials decreased delirium incidence, effectiveness of the separate components is unclear.

McAndrew et al. ^57^ applied quiet time from 1400 to 1600 among 72 mechanically ventilated ICU patients. In the 24 hours after starting quiet time, there was no increase in delirium rate and 19% of delirious patients improved to a negative CAM-ICU status. However, there was no significant effect on delirium in their analysis. Quiet time did lead to moderately improved sleep quality and less frequently administered sedatives which helped remove patients from mechanical ventilation. A pre-post research by Patel et al. ^7^ studied a nocturnal multidisciplinary environmental sleep promotion program in 338 non-delirious, non-sedated ICU patients. Their program included nocturnal quiet time with earplugs, eye mask, patient orientation, early mobilization, and sedation targets. The study group showed significant reduction in delirium incidence (by 33% p < 0.001), and a decrease in delirium duration (3.4 ± 1.4 versus 1. 2 ± 0.9 days, p = 0.021). Sleep quality and night-time light and noise levels were also improved in the study group, however reported noise levels were still higher than the WHO limits ^17^. They additionally reported a significant association between sleep efficiency and lower risk of developing delirium (OR 0. 90, 95% CI 0.84–0.97). A larger pre-post study (n=300) by Kamdar et al. ^58^ initiated a multi-faceted sleep promotion protocol consisting of three additive stages: 1) nightly quiet time and realignment of circadian rhythm, 2) sleeping with earplugs, eye masks, and soothing music, and 3) pharmacological targets to reduce sedatives. They reported decreased delirium incidence (OR = 0.46, 95% CI 0.23-0.89, p = 0.02) and perceived night-time noise in the study group, but no improvements in sleep quality.

## DISCUSSION

In this scoping review, existing literature was searched for studies on the impact of environmental risk factors and interventions on delirium: 21 studies were retrieved reporting effects of environmental risk factors on delirium and 16 studies reported experiments on possible solutions to modify the environment. Small sample sizes, heterogeneous study methods, and inconsistent results among reviewed studies proved the need for expanding research on impacts of environmental risk factors and efficacy of mitigations related to delirium.

### Modifiable ICU environmental risk factors for delirium

It is well recognized that ICU environments with round-the-clock activities and a high-tech setting have a negative impact on patients’ experience and clinical outcomes due to excessive noise, light, and disturbed sleep and circadian rhythm ^13, 47, 48^.

#### Noise

The WHO set recommendations for hospitals not to exceed an average of 30 dBA or a maximum of 35 dBA in treatment areas (maximum of 40 dBA at night) ^17^. A 2016 study by Hu et al. found average sound levels of 62.8 dB, with a mean level of 59.6 dB between 0000-0700, when investigating sound in various ICUs ^78^. Consistently, five reviewed articles measuring ICU sound pressure with or without noise modification interventions reported levels exceeding the WHO recommendations ^14, 18–20^.

A 2009 WHO report set night-time noise guidelines and reported on relationships between night time noise, sleep, and health. According to the report, excessive night-time noise (above 35 dB) disturb sleep, provoke annoyance and agitation, reduce cognition, impair communication and comprehension of surroundings, and contribute to psychiatric disorders. The combination of sleep disruption, decreased cognitive function, and lowered comprehension of surroundings associated with high noise levels may contribute to acute confusion and delirium ^79, 80^. In our review, two of three observational studies investigating association between high noise levels and ICU delirium found no significant effect of high noise levels on delirium incidence ^19, 20^. This result is surprising as it has been suspected that noise levels exceeding a normal threshold have detrimental effects on patient recovery, especially with regard to sleep and mental status. It is worth considering the difficulty in assessing the true effect of high noise levels in these two studies. First, there is no available baseline research to compare delirium incidence in high noise level ICUs versus those with statistically lower decibel values. It is possible the threshold for adverse effects is lower or higher than the most recently investigated decibel levels. In addition, Knauert et al. ^19^ mentioned a limitation for their study was inadequate statistical power to detect differences in decibel level between patient comparisons. For the study by Johansson et al. ^20^, their results need to be taken in context of using a non-validated delirium diagnosis protocol.

#### Light

During the daytime, normal light intensity is around 10000 lx and recommended night-time light levels conducive to sleep are below 30 lx ^59^. Natural fluctuation of light levels throughout the day contributes to the natural sleep-wake cycle by triggering release and suppression of melatonin. Alteration of the sleep-wake cycle and lack of daylight schedule have been shown to be associated with psychiatric diseases including depression, dementia, and delirium ^59^. Day-time light levels in the ICU are below normal daylight levels and above the threshold for sleep disruption at night ^59^. In a study by Hu et al. light intensity was measured over 24 hours near windows, in the center of rooms, and at eye level of mechanically ventilated patients. Average light intensity at these locations were 425 lx, 191 lx, and 388 lx respectively over 24 hours and 84 lx, 103 lx, and 87 lx between 2401 and 0759 ^78^. Minimal variation in daytime and night-time light levels disrupts the natural sleep-cycle and may contribute to patients becoming unable to distinguish day from night.

Abnormal natural light cycles are cited in recent literature as a potential modifiable risk factor for delirium management ^59^. Seven studies analyzing the impact of natural light on delirium incidence suggest this element of the ICU lacks a definitive causative relationship with development of the condition. Most of these studies enrolled critically ill patients whose condition gives them a higher likelihood of having consistently closed eyes compared to the general hospital population. It should be considered for future research that these patients’ retinas may not receive the same strength light stimulus as other populations, suggesting the need for ICU-specific lighting strategies. For the two seasonal studies, one found delirium was diagnosed significantly more in the winter than summer ^29^, while the other found exhaustive evidence ruling out a link between delirium and pre-hospital photoperiod exposure year-round ^31^. These findings suggest there are factors aside from seasonal light exposure affecting delirium. Additionally, of the three studies with a positive correlation between exposure to natural daylight or season of admission, the two natural daylight studies had vague descriptions of their measurements of patient’s exposure to natural or artificial light ^13, 46^. It is hard to assess whether the patient could have received benefits when the proximity of the stimulus to the patient is unclear.

As with excessive noise levels, further research into abnormal natural lighting cycles is necessary to delineate any threshold for adverse effects to patients’ well-being.

#### Sleep

Similar to our findings regarding effects of noise and light levels on delirium, reviewed articles on sleep showed mixed results for both forms of measure (electronic sleep monitoring and subjective reports). Recent literature states sleep is disturbed in ICU patients regardless of delirium ^19, 41^, and this concern is supported by the fact that unmeasurable sleep was found in non-delirious patients in included PSG studies. It is hard to compare results of included wireless monitoring studies, since different methodologies were used for each study, with different devices, leads, and whether or not they followed American Academy of Sleep Medicine standards. Similarly, it is difficult to compare findings from objective sleep monitoring protocols and subjective survey methods, and these need separate consideration. A major concern with analyzing subject sleep quality in delirious patients is patients with an altered mental state and/or confusion may not answer consistently or truthfully, and measures must be taken to assess whether answers are a correct representation of their condition.

### Environmental solutions to prevent or manage delirium

#### Noise modification

The negative impact of patient exposure to noise led to several studies focusing on noise pollution in the clinical environment. Mitigated exposure to noise levels might promote patient outcomes and staff satisfaction ^57, 81^. Noise reduction or abatement strategies include architectural features, behavioral alterations, alarm optimization, earplugs, headphones, and noise cancelling devices. Whilst these strategies have been studied in relation with improved noise levels and sleep promotion (Supplementary Table 3), further research is required to make evidence-based recommendations for the effect of noise reduction on delirium prevention and treatment.

Implementing ICU designs with acoustic features such as sound absorbers, reversible drawers to open both inside and outside the room, or room designs with the ability to locate alarmed devices or transfer alarms away from the patient, might improve exposure to noise and benefit delirium management ^47, 82, 83^. Zaal et al. demonstrated a lower delirium duration by modifying ICU design with acoustic considerations, however there was no change in delirium incidence rate ^47^. These strategies require major renovation or early construction planning, and further research is required to confirm cost-effectiveness and clinical benefits.

Staff and family conversations and care-activities are significant sources of ICU noise pollution ^16, 62, 63^. Although behavioral modification might be ineffective as a single-component intervention ^64^, low-cost adjustments such as limited bedside conversation, lowered voices, clustered care-activities, minimized TV and overhead use and volume, using vibrating pagers, and visual noise-warning devices may be necessary to achieve better results in sound reduction ^7, 14, 62, 63, 65, 66^, sleep improvement ^63^, and decreased delirium ^14^ To be successful, continuous awareness, education of staff on the impact of excessive noise exposure, and routine monitoring of implemented strategies is crucial ^7^. Technologies that help staff and visitors recognize excessive noise might complement implementation of behavioral strategies. Visual noise-warning devices display colored warnings at higher levels of noise and can be an effective, sustained noise reduction strategy ^65, 66^. Use of noise-warning systems has a greater impact on reduction of ambient noise compared with peak noise levels ^65, 66^. This is likely a result of change in staff behavior after visual warning while having no effect on medical equipment or alarms.

Alarms are a significant source of ICU noise pollution ^16, 62^, and a large portion are considered false positives ^84^. Studies show modifying ICU alarms by lowering volume, optimizing device settings, and filtering false alarms may reduce disturbing alarm noise ^85–87^. Schlesinger and colleagues equipped wearable earbuds with a frequency-selective silencing device, which could successfully filter ICU alarms while allowing patients to hear and communicate effectively without experiencing negative consequences of audible alarms ^72^. Optimization of alarms was used as an element of a noise reduction bundle and sleep promotion studies of this scoping review ^7, 14, 58^.

Abating environmental noise by earplugs or headphones appears feasible and effective to reduce noise and improve sleep in the ICU ^48, 56, 67–70^. Here, one study failed to prove benefits of using earplugs and eye masks during sleep on delirium ^56^, while another earplug trial decreased risk of confusion, and delayed initiation of cognitive disturbances with no significant effect on incidence of delirium ^48^. Given the potential effectiveness and low costs, this method is frequently used in multi-component interventions ^7, 58^; however, non-compliancy is a major implementation issue in earplugs studies ^56^. A recent meta analysis ^81^ reported a 13.1% (95% CI, 7.8–25.4) rate of non-compliancy due to intolerance, anxiety, or accidental removal of earplugs. Headphones with active noise cancellation technology might improve patient outcomes by mitigating exposure to noise. Gallacher et al. modeled an experiment by embedding sound meters in the auditory meatus of polystyrene model heads located near patients’ beds in a cardiac ICU ^70^. They demonstrated a significant reduction in overall noise exposure and exposure to high intensity noises using noise cancelling headphones.

Despite inconsistent results of the reviewed studies on efficacy of noise modifications on delirium, this review suggests considering physical design features and multi-component noise reduction programs may benefit delirium prevention or management. This is consistent with current recommendations suggesting multi-component interventions to achieve adequate noise reduction ^81^; Van de Pol et al. ^14^ reduced delirium incidence by implementing a noise reduction program consisting of behavioral strategies, device optimization, and earplugs. However, there is a need for high-quality randomized control trials with larger sample sizes to evaluate efficacy, sustainability, and long-term effects of noise modification interventions with a focus on delirium.

#### Light modification

Optimized circadian rhythm needs bright days and dark nights. Various light modification strategies have been proposed to follow circadian rhythms. These are categorized as such: decreasing night-time light exposure, and increasing daylight.

Round-the-clock ICU activities make nigh-time light reduction challenging to maintain a level of light sufficient for providing care, but not disturbing sleep. Dimming lights as part of quiet time strategies is effective to mitigate intensity of light during quiet time hours, however, this may cause variation in perceived light and consequently cause sleep disturbance ^87^. Possible solutions are clustering care-activities to reduce bedside interruptions ^7^ and use of portable lighting pods with less blue wavelength during the night ^88^. Whilst the trial of sleep mask and earplugs by Demoule at al. ^56^ failed to show benefit to delirium, eye masks are effective in promoting sleep by light abatement ^7, 74, 76, 77^. However, poor compliance using eye masks due to accidental removal, or anxiety/claustrophobia, and the risk of sensory deprivation in mechanically ventilated patients, remains challenging ^56^.

Environmental modification to increase daylight exposure is possible through architectural considerations of promoting natural lighting or utilizing artificial illumination. Research into whether windows allow enough light to promote sleep-wake cycles and prevent delirium, and whether seasonal light levels contribute to delirium, has been conducted with inconclusive findings ^30, 32, 73^. From our results, the greatest interventional effect on delirium was from bright light therapy.

Our review included five studies on BLT, three reporting a significant effect on delirium incidence or severity ^51, 53, 55^ with sleep promoted in four studies ^52–55^. BLT has the greatest effect between 2500 and 10000 lx for 30 to 60 minutes, with a shorter duration for greater intensities of light, when administered either at twilight or dawn to obtain a circadian effect ^60^. The BLT in this review applied 2000-10000 lx of illuminance for between one and four hours. The use of 2000 lx was effective in improving sleep quantity and functional status during management of delirium as part of a bundle. The use of 5000 lx was associated with decreased delirium incidence in two of three studies and the use of 10000 lx, as an adjunctive treatment with risperidone, was associated with a decrease in delirium severity ^53^. While BLT may help regulate sleep-wake cycles and prevent/treat delirium, research into melatonin secretion and circadian rhythms suggests periods of darkness play as large a role as daytime light levels in promoting sleep and preventing delirium ^59, 89^. The importance of light and darkness prompts a need for research into effects of dynamic lighting systems. This review included three studies focused on dynamic lighting among sedated and non-sedated patients, using lighting systems which produced cooler blue light in the mornings and shifted towards warmer tones as the day progressed. The lighting systems produced different levels of intensity throughout the day, reaching a peak of between 750 and 4000 lx and a minimum level of 0 lx. None of these studies showed significant effects on delirium ^15, 49, 50^; however, they used peak light levels below normal daytime levels.

Maintaining a circadian rhythm, by nocturnal darkness and BLT, as a low-cost, low-risk, easy-to-apply intervention can help improve patient outcomes. Research is required to investigate the use of dynamic lighting with higher peak light intensities or the combination of dynamic lighting and BLT. Additionally, there is a need for defining effective characteristics of light modification strategies for sedated and non-sedated patients. Sedated patients may have disrupted circadian rhythm of melatonin ^90^, and application of light therapies might have limited retina stimuli when eyes are closed. Studies comparing efficacy of light modifications on prevention or treatment of delirium among these two groups of patients, with application of different intensity levels of light in closed-eyes patients, might be of benefit.

#### Intervention bundles (light and noise modification)

There is a growing interest in using quiet time interventions to promote sleep. Quiet time is defined as a designated period of time in which there is a limitation in bedside interruptions and a reduction in noise and light levels. Quiet time protocols have successfully reduced sound pressure, improved sleep quality, and reduced the use of sedatives ^87, 91–94^, but effects of quiet time on delirium development needs further research. McAndrew et al. implemented a daily quiet time protocol in ICU patients and reported inconclusive results on delirium scores and moderate improvement in sleep perception ^57^. Two neurocritical ICU studies have implemented a two hour quiet time during day and night ^93, 94^A significant improvement in subjective sleep and increased staff satisfaction was achieved ^93, 94^. They reported decreased light by 75-85% and noise by 15%, with results being more significant during day-shift quiet time; this might be due to overall lower levels of nocturnal light and noise ^93^.

Sleep promotion protocols utilize noise and light control strategies with other components, such as patient orientation, early mobilization, medication optimization, and sedation targets to improve sleep in quality and quantity. Here we included two sleep promotion studies reporting results on delirium, however future research is needed to evaluate which component of sleep promotions are effective in reducing delirium. Patel et al. ^7^ significantly improved sleep quality and reduced delirium incidence by implementing a non-pharmacological multidisciplinary sleep program. They raised protocol compliance to > 90% by ongoing education, signage and posters, monitoring, and spot-checking program quality by experienced nurse champions. Interestingly, a large sleep promotion study by Kamdar et al., decreased delirium incidence while there was no effect on sleep ^58^. It is not clear if improvements in delirium are attributable to sleep, emphasizing the need for future studies focused on single interventions or single components of multifaceted interventions with regard to delirium results.

The main strength of this review is synthesizing results of both observational association studies and interventional studies. This approach details a broader picture of the current state of this research field and bridges the gap between establishing correlational relationships and continuation of experimental trials. A major limitation of this review is the narrow search method. By searching one database (Pubmed) and the included articles’ reference lists, there is likely additional literature available to expand our findings, however the authors did a hand search within related journals, Embase, and Google Scholar databases to include existing interventional research articles. Another limitation was generated data from reviewed studies did not have full details, and quality of evidence was not evaluated among studies; however, this review was intended to be a literature mapping with limited description of relevant publications.

## CONCLUSION

This review of studies investigating the association between delirium and either high noise levels, abnormal amounts of natural daylight, and/or sleep disruptions did not reveal a clear relationship between delirium and these variables. It is recommended to perform additional research into more comprehensive, but related, risk factors to find a stronger predictor. Additional research could include analyses of specific noise sources or a comparison between overcast, rainy, and sunny times. This study demonstrated overall evidence supporting effectiveness of environmental interventions on delirium is of low confidence. Current literature lacks randomized control trials with larger sample sizes to evaluate efficacy of intervention on delirium and its long-term outcomes. Another knowledge gap is lack of adequate conclusive research on single-component interventions. The interventional bundle studies lead to uncertainty about which component impacts the result. Given the low-cost and non-invasive nature of environmental modifications and their potential beneficial role in reduction of modifiable risk factors, it is recommended to implement these interventions in current practice, especially as multi-component bundles.

## Data Availability

NA

